# Cohort profile: St. Michael’s Hospital Tuberculosis Database (SMH-TB), a retrospective cohort of electronic health record data and variables extracted using natural language processing

**DOI:** 10.1101/2020.09.11.20192419

**Authors:** David Landsman, Ahmed Abdelbasit, Christine Wang, Michael Guerzhoy, Ujash Joshi, Shaun Mathew, Chloe Pou-Prom, David Dai, Victoria Pequegnat, Joshua Murray, Kamalprit Chokar, Michaelia Banning, Muhammad Mamdani, Sharmistha Mishra, Jane Batt

**Author notes:** These authors contributed equally to this work (Sharmistha Mishra, Jane Batt). Corresponding author: Jane Batt, MD, PhD.

## Abstract

**Background:** Tuberculosis (TB) is a major cause of death worldwide. TB research draws heavily on clinical cohorts which can be generated using electronic health records (EHR), but granular information extracted from unstructured EHR data is limited. The St. Michael’s Hospital TB database (SMHTB) was established to address gaps in EHR-derived TB clinical cohorts and provide researchers and clinicians with detailed, granular data related to TB management and treatment.

**Methods:** We collected and validated multiple layers of EHR data from the TB outpatient clinic at St. Michael’s Hospital, Toronto, Ontario, Canada to generate the SMH-TB database. SMH-TB contains structured data directly from the EHR, and variables generated using natural language processing (NLP) by extracting relevant information from free-text within clinic, radiology, and other notes. NLP performance was assessed using recall, precision and F_1_ score averaged across variable labels. We present characteristics of the cohort population using binomial proportions and 95% confidence intervals (CI), with and without adjusting for NLP misclassification errors.

**Results:** SMH-TB currently contains retrospective patient data spanning 2011 to 2018, for a total of 3298 patients (N=3237 with at least 1 associated dictation). Performance of TB diagnosis and medication NLP rulesets surpasses 93% in recall, precision and F_1_ metrics, indicating good generalizability. We estimated 20% (95% CI: 18.4-21.2%) were diagnosed with active TB and 46% (95% CI: 43.8-47.2%) were diagnosed with latent TB. After adjusting for potential misclassification, the proportion of patients diagnosed with active and latent TB was 18% (95% CI: 16.8-19.7%) and 40% (95% CI: 37.8-41.6%) respectively

**Conclusion:** SMH-TB is a unique database that includes a breadth of structured data derived from structured and unstructured EHR data. The data are available for a variety of research applications, such as clinical epidemiology, quality improvement and mathematical modelling studies.

## Introduction

Tuberculosis (TB) is the top infectious killer worldwide, resulting in 1.6 million deaths in 2017 (1). 1.7 billion people carry the latent form of the infection, of whom 10% at minimum, will develop the active, infectious form of disease. Latent TB infection (LTBI) progression to active disease can be prevented and TB can be cured, with appropriate antibiotics taken over many months. TB is endemic in many low-income countries and particularly prevalent in Asia and Africa. The World Health Organization recommends the treatment of LTBI as part of the global “End TB Strategy”, and an achievable goal critical to TB elimination in high-income countries (2,3).

Given the burden of active TB disease is disproportionately carried in low-resource settings, research addressing disease epidemiology, treatment (including clinical trials and programs of delivery), and the use and utility of innovative and point of care diagnostics is often completed in the populations of countries with highest burden of TB. The prevalence of LTBI on the other hand, is considerable even in high-income countries (CDC estimates 13,000,000 people living the USA have LTBI (4)) and thus research ranging from basic pathogenesis to program development can be conducted on the global population. Indeed while advances in biomedical research over the past 1 to 2 decades have delivered successes ranging from rapid point-of-care diagnostics testing for pulmonary TB to the development of novel therapeutics such as bedaquiline and delamanid, many questions remain, including, for example, discovering biomarkers that precisely indicate individuals at risk of LTBI activation and developing programs of TB care that ensure efficacy, are equitable and resilient (1,5).

Many primary care practices and hospitals in high-income countries have curated electronic health record (EHR) data for research and surveillance (6–9), that improve ease of access to information and data sharing for collaborative work. The use of EHRs in hospital and office-based clinical practices has risen substantially in the past decade, providing rich data sources that have the potential to simultaneously improve patient care and advance research initiatives (10,11). Most EHR-derived databases are however limited to structured data, such as demographic information collected at patient registration, laboratory tests and results and diagnostic codes used in physician billing. As such, the rich, granular data embedded within unstructured (text) data from dictated notes on both hospital admitted and clinic patients are excluded (12,13) unless these variables are abstracted via manual chart review (14,15) or natural language processing (NLP) (16–18). We developed the first digital retrospective clinical database that combines structured data, unstructured (text) data, and variables derived from transforming unstructured data to structured data using natural language rulesets, among patients assessed in an inner-city outpatient TB clinic at St Michaels Hospital (SMH) of Unity Health Toronto in Toronto, Ontario, Canada.

Approximately 2000 people (5.6 per 100,000 people) are diagnosed with active TB in Canada (19) annually and 1.3 million are estimated to have LTBI. The SMH TB clinic cares exclusively for individuals with suspected or diagnosed active TB and LTBI, seeing 1800-2200 patient-visits each year, and assessing and developing a diagnostic and management plan for 670-800 new patients each year.

In this paper we describe the SMH-TB database, which aims to be a resource for scientists who are conducting research into many facets of TB, ranging from observational epidemiology to emulated trials and quality improvement and implementation science research. The purpose of this profile is to describe our methodology, present the cohort and the database validation. Access to the database is available to collaborators wishing to work with the research team of the SMH TB clinic. The NLP rulesets developed to extract variables from the unstructured data in the EHR are publicly available on GitHub (20).

## Materials and methods

### Cohort Description

The database compiles all data available on all TB clinic patients (N=3298) treated at SMH from April 2011 to December 2018. The database contains socio-demographic information surrounding immigration, housing status, and insurance, and clinical information including laboratory and imaging results, co-morbidities, diagnoses and treatment. Ethics approval for development and validation of the database was obtained from the Unity Health Toronto Research Ethics Board (REB 19-080). Patient consent was not required or obtained as per the Tri-Council Policy Statement 2 (TCPS2), since only retrospective data were collected from clinical charts (21).

Patients are referred to the TB outpatient clinic predominantly from Public Health Units in the Greater Toronto area (population of 6 million), Canada Immigration and Citizenship, Occupational Health and Safety Departments of Toronto area hospitals, community health care professionals (physicians, nurse-practitioners), and SMH staff physicians caring for an admitted patient or a patient in the emergency room (ER). When including a patient in our database we consider all available encounters, including inpatient admissions and ER visits.

### Data Collection

St. Michael’s Hospital EHR is managed by several systems. The Enterprise Data Warehouse (EDW) stores and manages structured data including patient demographics and medical test results. Soarian stores the unstructured patient data, which includes dictated clinical notes. SMHTB retrieved data of patients registered and assessed in the TB outpatient clinic to provide a comprehensive description of patient characteristics, disease, management and clinical trajectory.

SMH-TB is restricted to a start-date of April 2011, which is the date of initiation of EHR at SMH. Fig 1 shows the data flow and data sources for the SMH-TB database.

**Fig 1:**
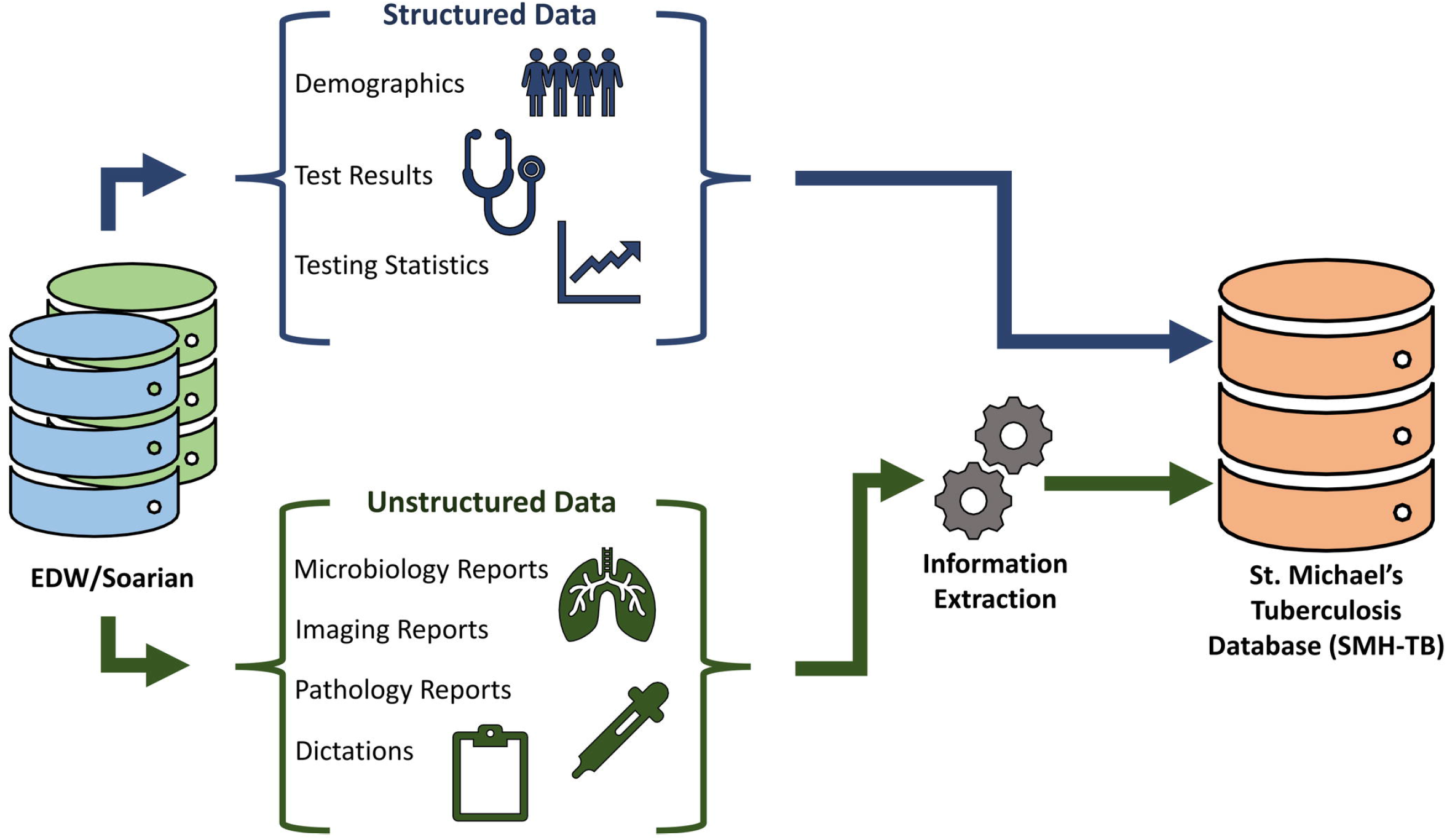
Data sources for SMH-TB Database.

The SMH-TB database stores patient characteristics and encounter data in separate tables, which can be linked together using unique, de-identified patient or encounter IDs. Fig 2 presents the tables provided in SMH-TB, and the granularity of the data they contain. A detailed collection of all the variables available in the database is provided in Table 1.

**Fig 2:**
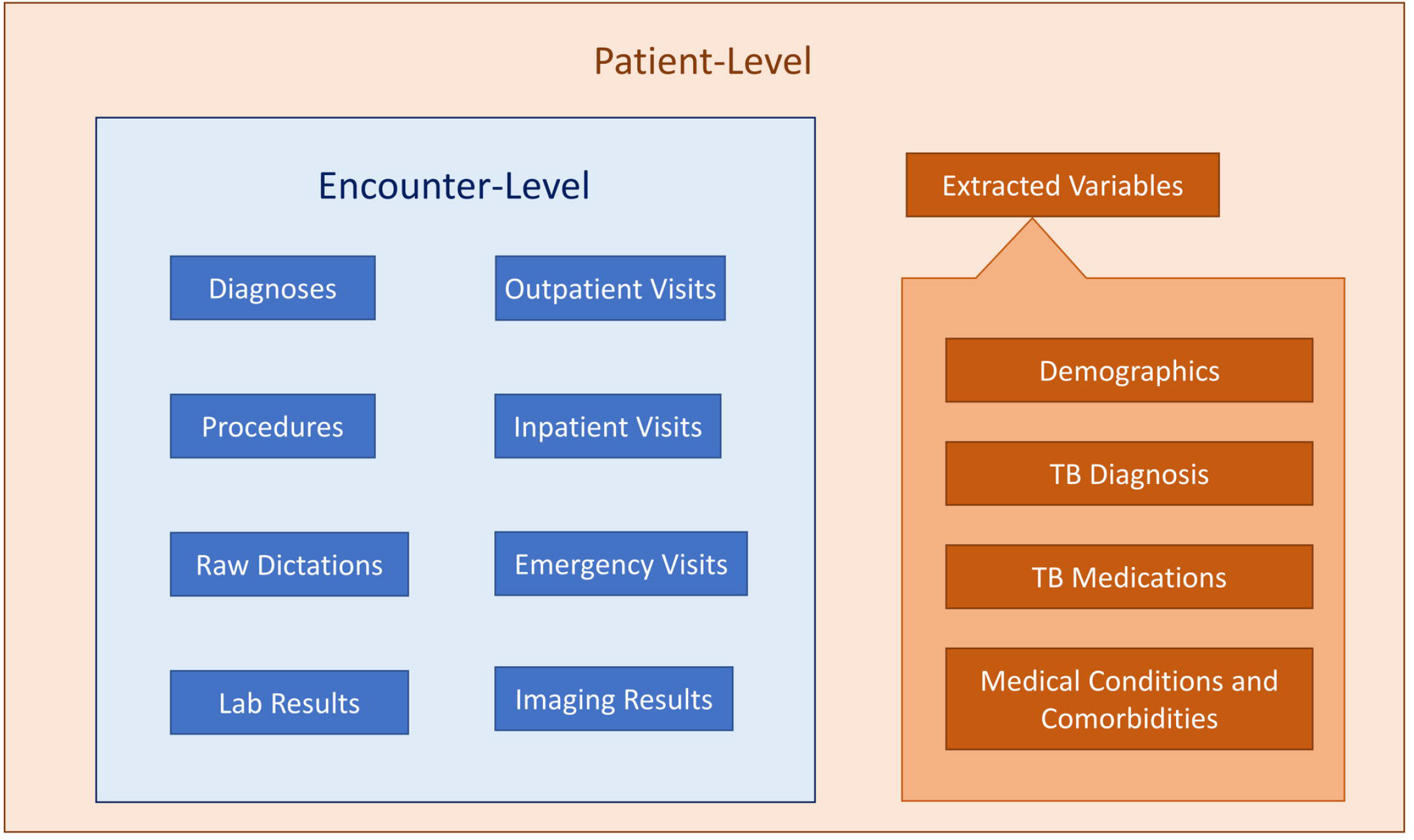
Patient-level and Encounter-level Data in SMH-TB.

**Table 1:** Variables available in SMH-TB from both structured and unstructured sources.

|  |  |
| --- | --- |
| Demographics | Tuberculosis Diagnosis |
| Patient ID | Known TB exposure* |
| MRN | BCG vaccination status* |
| Sex | TST performed* |
| Date of birth | TST induration* |
| Street address | TST interpretation* |
| Postal code<sup>a</sup> | IGRA performed* |
| Country of origin* | IGRA interpretation* |
| Year of immigration* | Diagnosis of active TB* |
| Immigration status | Diagnosis of LTBI* |
| Housing status |  |
| Insurance status | **Tuberculosis Medications** |
| Patient is a healthcare worker* | Ever started isoniazid* |
|  | Ever started rifampin* |
| Encounter Details | Ever started pyrazinamide* |
| Encounter ID | Ever started ethionamide* |
| Encounter type | Ever started vitamin B6* |
| Encounter date |  |
| Direct cost<sup>b</sup> | **Medical Conditions and Comorbidities** |
| Indirect cost<sup>c</sup> | Autoimmune conditions<sup>d*</sup> |
|  | Diabetes* |
| **Aggregate Variables** | Hematological malignancy* |
| Number of sputum inductions | Non-hematological malignancy* |
| Number of chest x-rays | Transplant performed* |
| Number of chest computed tomography | Renal failure<sup>e*</sup> |
| Hospital admission during course of TB outpatient care | Silicosis* |
| Number of emergency room visits during course of TB outpatient care | Hepatitis B |
|  | Hepatitis C |
| **Laboratory Results** | HIV status* |
| AST |  |
| ALT | **Microbiology Reports\*\*** |
| CBC (Hb, Platelets, WBC) | **Radiology Reports\*\*** |
| Cr | **Pathology Reports\*\*** |
| Bilirubin |  |

MRN: Medical record number; AST: Aspartate transaminase; ALT: Alanine transaminase; CBC:
Complete blood count; Hb: Hemoglobin; WBC: White blood cells; Cr: Creatinine; TB: Tuberculosis;
BCG: Bacillus Calmette–Guérin; TST: Tuberculin sensitivity test; IGRA: Interferon gamma release
assay; LTBI: Latent tuberculosis infection; HIV: Human immunodeficiency viruses
<sup>a</sup>The database only stores the Forward Sortation Area portion of the postal code of the patient's residence.
<sup>b</sup>Direct cost corresponds to health care services directly associated with the patient's care including all nursing, allied health, diagnostic and therapeutic services, pharmaceutical and medical/surgical supplies for each visit.
<sup>c</sup>Indirect cost corresponds to administrative and support services performed on behalf of all patients including information system and housekeeping overheads.
<sup>d</sup>Autoimmune conditions include: Sjogren's syndrome, arthropathy, spondyloarthropathy, psoriatic arthritis, rheumatoid arthritis, reactive arthritis, mixed connective tissue disease, connective tissue disease, systemic lupus erythematosus, CREST syndrome, dermatomyositis, Wegener's granulomatosis, Goodpasture syndrome, vasculitis and psoriasis.
<sup>e</sup>Renal failure includes: nephropathy, renal insufficiency and glomerulonephritis.
\*Variables collected from unstructured dictations and reports using natural language rulesets

#### Removing identifiable information

There are two versions of SMH-TB. The full version includes indelible patient identifiers such as a patient’s provincial health insurance (Ontario Health Insurance Plan) number; their SMH-specific medical record number; all patient encounters whose encounter record is specific to a given patient; laboratory test records whose encounter record is also specific to a given patient; and all unstructured text data per encounter per patient. The patient identifiers allow for a fully linked database, which can be updated and linked via future data extraction. The identifiable unstructured data are also retained to support the development and testing of additional natural language rulesets.

The de-identified version of SMH-TB is the version that will be primarily used for research studies. It excludes the unstructured data and has been stripped of the following: hospital patient ID, hospital encounter ID, address and day and month of date of birth. Each patient and encounter is then re-coded with new unique IDs, and with the age in years on the date of the first TB clinic encounter

#### Patient identification and validation

The Decision Support Services (DSS) at SMH identified encounters which were coded as services provided in the TB outpatient clinic to identify all TB patients. We then randomly selected a list of 200 patients seen in the TB outpatient clinic (using clinic schedules with unique patient identifiers stored separately from the EDW) to manually validate the codes used by DSS to identify TB clinic outpatients, and validated that all (100%) identified patients were registered in the TB clinic. To ensure high specificity of our identification of TB clinic patients, we examined additional metadata (such as a mention of the TB clinic in the patient’s dictations) and removed patients without matching metadata. SMH-TB therefore may include the rare patient where the clinic visit codes in the EDW erroneously labelled a visit as a TB clinic visit, but this estimate is expected to be <0.2% because of the additional metadata checks. The hospital unique patient identifier for each individual was then cross referenced to lists of all individuals with inpatient stays and ER visits to derive TB patient data from all sites of contact for TB care.

#### Data transformation (unstructured text to structured variables)

Unstructured clinician dictations were used to create patient-level variables on demographics, TB diagnosis, TB medications and comorbidities. The data for these variables were extracted using rule-based information extraction tool CHARTextract (22). CHARTextract uses regular expressions in order to perform pattern matching on text. Regular expressions have been used to perform data extraction and even classification due to their high expressivity (17,23,24). These capabilities come at the cost of a complex syntax, and thus rule creation typically involves the expertise of a clinician who understands the subject matter and an interpreter who can express the idea into regular expression syntax. We created a tiered rule system, where primary rules are used to filter text at the sentence level using a scoring system and secondary rules can be used to further enhance the weighting of the sentence. The tool applies the user-created rules to the data and extracts the variables on-the-fly. The interface displays mismatches between the tool prediction and the gold-standard label. Users can iterate on the rule creation process, allowing for easy refinement and quick development of the rules. Fig 3 shows a component of a ruleset for extracting diagnosis of active tuberculosis.

**Fig 3:**
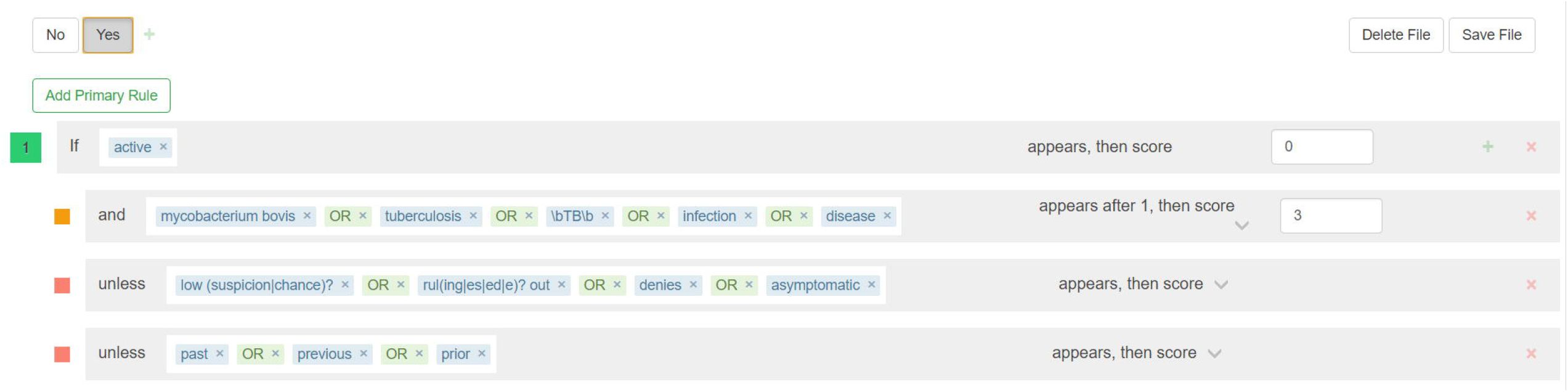
Example of a component of a ruleset for extracting a variable (active TB diagnosis) from unstructured text in clinical dictations (using CHARTextract)

In order to create the rulesets used by CHARTextract, two clinicians (JB, SM) from the TB outpatient clinic were consulted on dictation language and style. Clinicians (JB, SM, AA, and KC) and a medical student (CW) manually labeled dictations for 200 patients from a subset of the dataset to be used for validation. The set of 200 patients was selected from consecutive clinic visits based on registered patient lists external to the EHR. This was done using the QuickLabel tool which provides a user interface for streamlined labelling of specific variables, as well as the option to label multiple variables simultaneously (25). Refinement of the natural language rulesets was done by comparing the labels extracted by the rulesets via CHARTextract with the manual labels. The refined rulesets are available as a real-time source as additional variables from unstructured data (microbiology, radiology, and pathology reports) are generated (20).

#### Evaluation of data extraction

To measure the performance of our rulesets and evaluate their generalizability to unseen data, we calculated accuracy, recall, precision and F_1_ scores. Recall (sensitivity) measures the ability of the classifier to correctly distinguish true positive from false negative examples. Precision (positive predictive value) measures the ability of the classifier to correctly distinguish true positive from false positive examples. The F_1_ score computes a harmonic mean of precision and recall. Recall, precision and F_1_ score were averaged across variable labels.

#### Binomial proportions estimated from extracted variables

We used the refined rulesets to extract variables from the full dataset of patients with at least 1 dictation (N=3237). We converted “Yes/No/Not recorded” and “Positive/Negative/Unknown/Not recorded” variables into binary 0-1 variables by assigning a value of 1 to patients with an extracted value of “Yes” or “Positive”, and a value of 0 otherwise. We estimated the proportion and 95% confidence intervals of patients for which the rulesets extracted “Yes” or “Positive” for these variables using two methods: (1) logistic regression model without covariates, and (2) MCSIMEX model that accounts for the misclassification error in the extracted variables that was calculated from the set of 200 manually abstracted patients (26). Briefly, for a binary random variable Y, we estimate the probability P(Y=1) using a logistic regression model without covariates, given by:

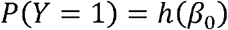

where h is the logistic function. Under the MC-SIMEX model, the binary random variable was observed with misclassification errors, denoted by *Y*\*. We estimate the probability P(*Y*\*=1)as:

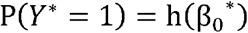

where β_0_^*^ is defined as:

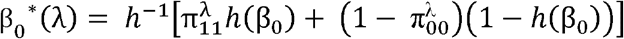

Π_00_ and π_11_ denote the specificity and sensitivity of *Y*\*, respectively, and λ is the misclassification parameter. The final estimate for β_0_^*^ is computed by a simulation-extrapolation procedure described in (26).

## Results

### Population

A patient overview based on demographics is presented in Table 2. 3298 patients were included in the database. The median age of the patients is 45 years, with an interquartile range of 34 to 58. There is a higher percentage of females than males in the cohort, around 57%. At least 79% of the clinic patients were born outside of Canada, based on data extracted from patients’ dictations. The vast majority of patients were adequately housed, with publicly funded provincial health care insurance (OHIP).

**Table 2:**
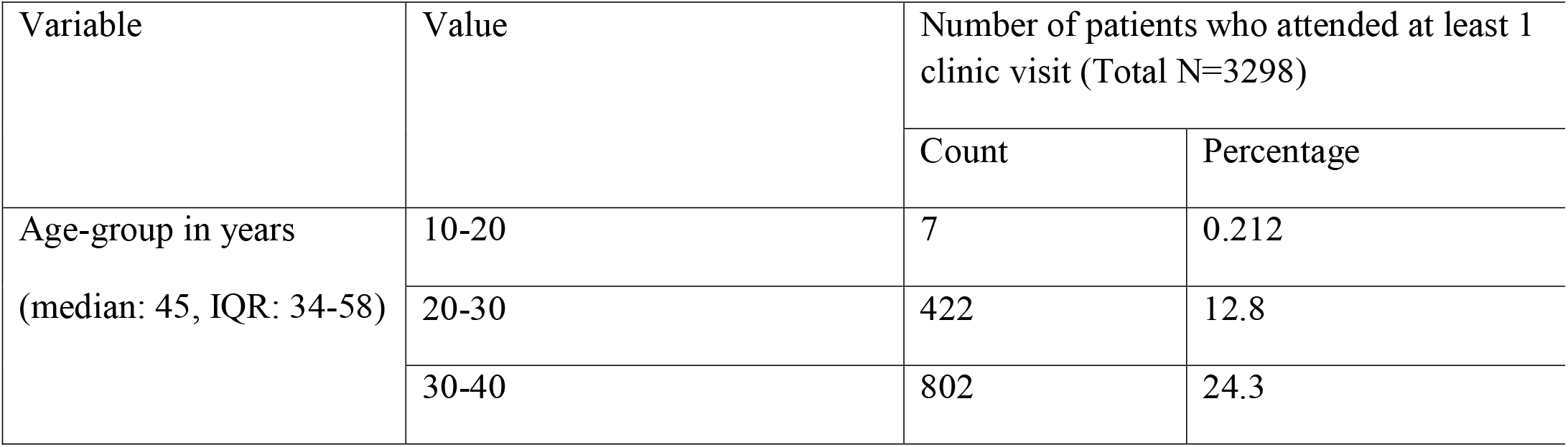
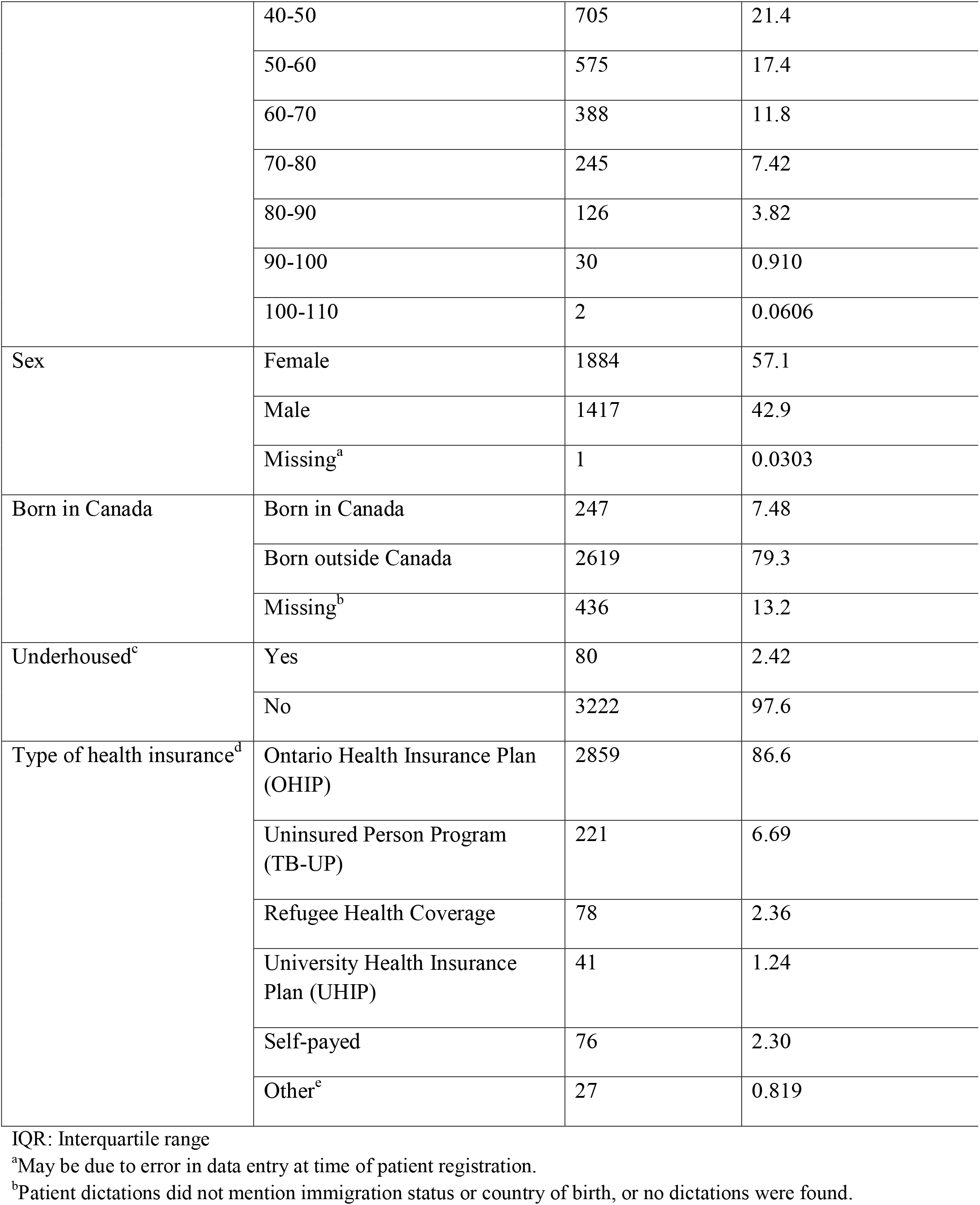
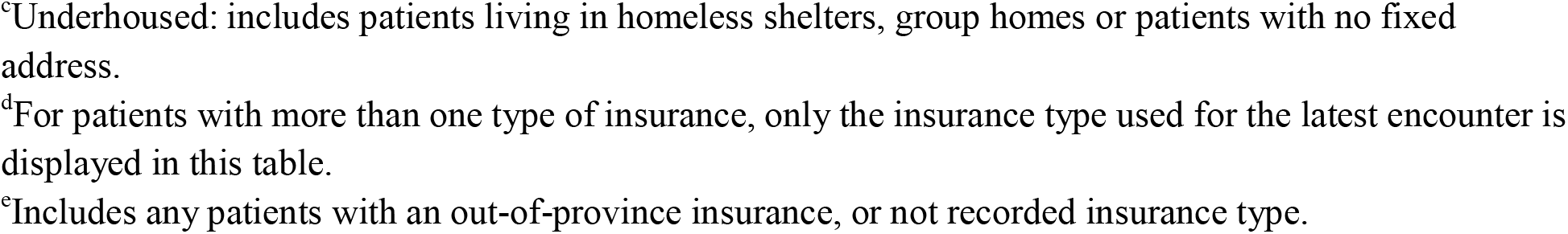
Demographics of the patients included in the SMH-TB database, 2011-2018.

| Variable | Value | Number of patients who attended at least 1 clinic visit (Total N=3298) |  |
| --- | --- | --- | --- |
|  |  | Count | Percentage |
| Age-group in years<br>(median: 45, IQR: 34-58) | 10-20 | 7 | 0.212 |
|  | 20-30 | 422 | 12.8 |
|  | 30-40 | 802 | 24.3 |
|  | 40-50 | 705 | 21.4 |
|  | 50-60 | 575 | 17.4 |
|  | 60-70 | 388 | 11.8 |
|  | 70-80 | 245 | 7.42 |
|  | 80-90 | 126 | 3.82 |
|  | 90-100 | 30 | 0.910 |
|  | 100-110 | 2 | 0.0606 |
| Sex | Female | 1884 | 57.1 |
|  | Male | 1417 | 42.9 |
|  | Missing<sup>a</sup> | 1 | 0.0303 |
| Born in Canada | Born in Canada | 247 | 7.48 |
|  | Born outside Canada | 2619 | 79.3 |
|  | Missing<sup>b</sup> | 436 | 13.2 |
| Underhoused<sup>c</sup> | Yes | 80 | 2.42 |
|  | No | 3222 | 97.6 |
| Type of health insurance<sup>d</sup> | Ontario Health Insurance Plan (OHIP) | 2859 | 86.6 |
|  | Uninsured Person Program (TB-UP) | 221 | 6.69 |
|  | Refugee Health Coverage | 78 | 2.36 |
|  | University Health Insurance Plan (UHIP) | 41 | 1.24 |
|  | Self-paid | 76 | 2.30 |
|  | Other<sup>e</sup> | 27 | 0.819 |

IQR: Interquartile range
<sup>a</sup>May be due to error in data entry at time of patient registration.
<sup>b</sup>Patient dictations did not mention immigration status or country of birth, or no dictations were found.
<sup>c</sup>Underhoused: includes patients living in homeless shelters, group homes or patients with no fixed address.
<sup>d</sup>For patients with more than one type of insurance, only the insurance type used for the latest encounter is displayed in this table.
<sup>e</sup>Includes any patients with an out-of-province insurance, or not recorded insurance type.

### Evaluation of data extraction

A summary of the rulesets’ performance metrics for the 25 variables extracted from unstructured dictations is presented in Table 3. Diagnosis of active TB and LTBI rulesets had 97.5% and 96% accuracy, and 97.4% and 94.7% F_1_ score, respectively. Rulesets for extracting TB medications generally achieved above 90% accuracy, recall and precision metrics.

**Table 3:**
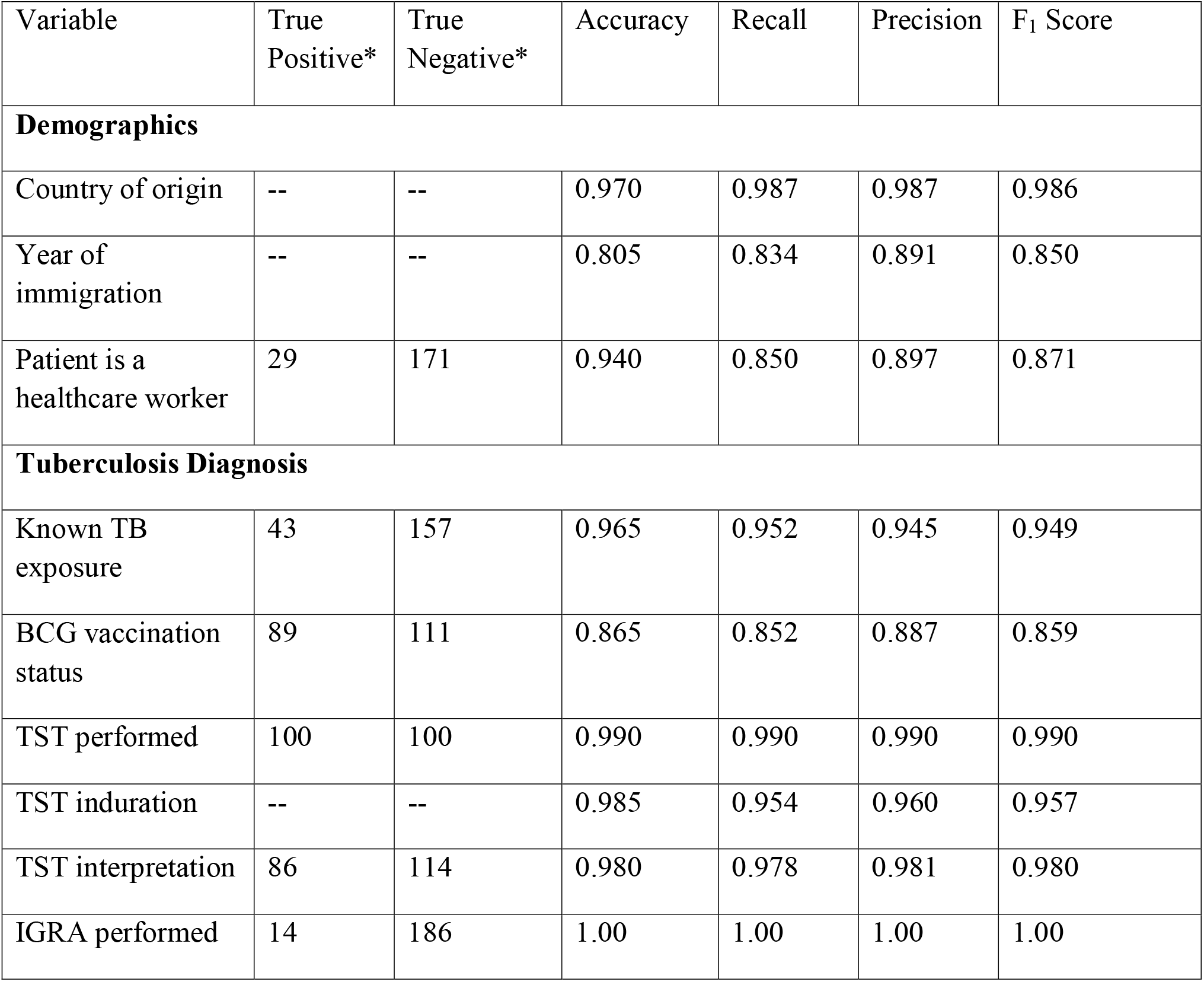
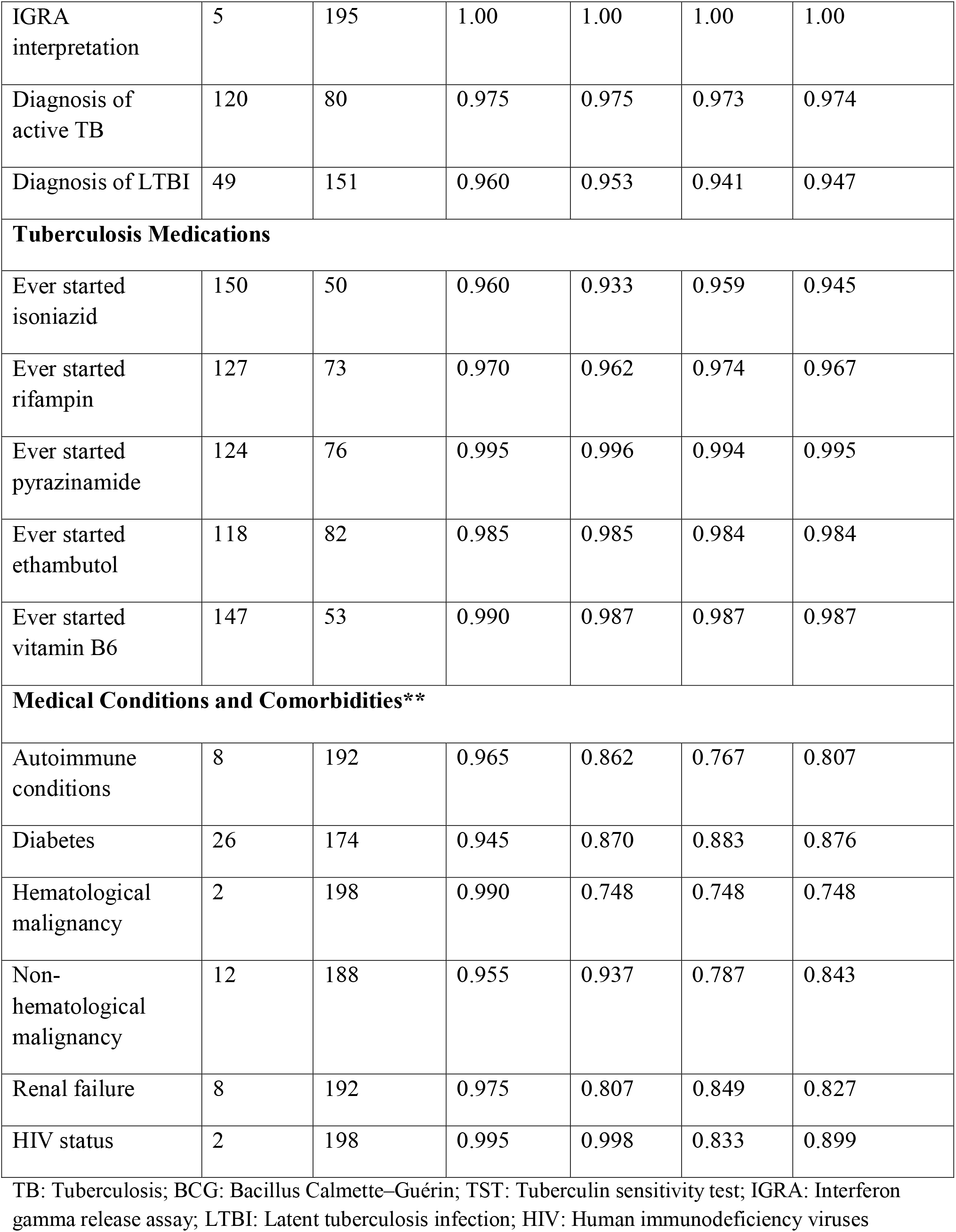
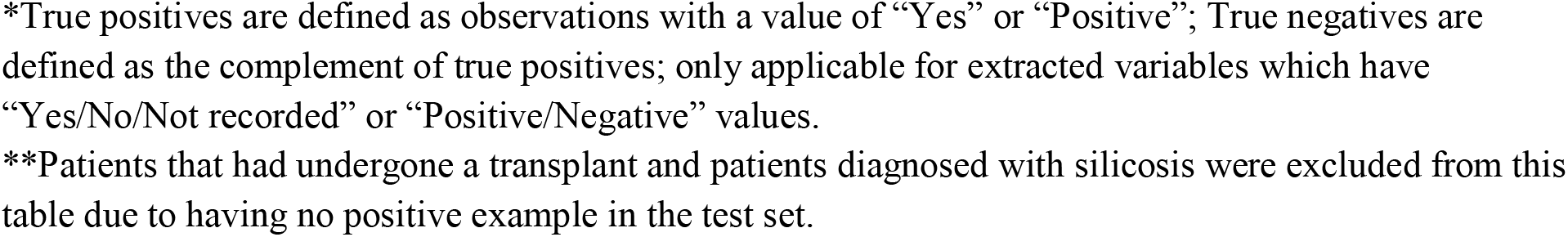
Summary of performance metrics on test set for variables extracted from unstructured dictations. Patients included in test set: N = 200.

| Variable | True Positive* | True Negative* | Accuracy | Recall | Precision | F<sub>1</sub> Score |
| --- | --- | --- | --- | --- | --- | --- |
| **Demographics** |  |  |  |  |  |  |
| Country of origin | -- | -- | 0.970 | 0.987 | 0.987 | 0.986 |
| Year of immigration | -- | -- | 0.805 | 0.834 | 0.891 | 0.850 |
| Patient is a healthcare worker | 29 | 171 | 0.940 | 0.850 | 0.897 | 0.871 |
| **Tuberculosis Diagnosis** |  |  |  |  |  |  |
| Known TB exposure | 43 | 157 | 0.965 | 0.952 | 0.945 | 0.949 |
| BCG vaccination status | 89 | 111 | 0.865 | 0.852 | 0.887 | 0.859 |
| TST performed | 100 | 100 | 0.990 | 0.990 | 0.990 | 0.990 |
| TST induration | -- | -- | 0.985 | 0.954 | 0.960 | 0.957 |
| TST interpretation | 86 | 114 | 0.980 | 0.978 | 0.981 | 0.980 |
| IGRA performed | 14 | 186 | 1.00 | 1.00 | 1.00 | 1.00 |
| IGRA interpretation | 5 | 195 | 1.00 | 1.00 | 1.00 | 1.00 |
| Diagnosis of active TB | 120 | 80 | 0.975 | 0.975 | 0.973 | 0.974 |
| Diagnosis of LTBI | 49 | 151 | 0.960 | 0.953 | 0.941 | 0.947 |
| **Tuberculosis Medications** |  |  |  |  |  |  |
| Ever started isoniazid | 150 | 50 | 0.960 | 0.933 | 0.959 | 0.945 |
| Ever started rifampin | 127 | 73 | 0.970 | 0.962 | 0.974 | 0.967 |
| Ever started pyrazinamide | 124 | 76 | 0.995 | 0.996 | 0.994 | 0.995 |
| Ever started ethambutol | 118 | 82 | 0.985 | 0.985 | 0.984 | 0.984 |
| Ever started vitamin B6 | 147 | 53 | 0.990 | 0.987 | 0.987 | 0.987 |
| **Medical Conditions and Comorbidities\*\*** |  |  |  |  |  |  |
| Autoimmune conditions | 8 | 192 | 0.965 | 0.862 | 0.767 | 0.807 |
| Diabetes | 26 | 174 | 0.945 | 0.870 | 0.883 | 0.876 |
| Hematological malignancy | 2 | 198 | 0.990 | 0.748 | 0.748 | 0.748 |
| Non-hematological malignancy | 12 | 188 | 0.955 | 0.937 | 0.787 | 0.843 |
| Renal failure | 8 | 192 | 0.975 | 0.807 | 0.849 | 0.827 |
| HIV status | 2 | 198 | 0.995 | 0.998 | 0.833 | 0.899 |

TB: Tuberculosis; BCG: Bacillus Calmette–Guérin; TST: Tuberculin sensitivity test; IGRA: Interferon gamma release assay; LTBI: Latent tuberculosis infection; HIV: Human immunodeficiency viruses
\*True positives are defined as observations with a value of “Yes” or “Positive”; True negatives are defined as the complement of true positives; only applicable for extracted variables which have “Yes/No/Not recorded” or “Positive/Negative” values.
\*\*Patients that had undergone a transplant and patients diagnosed with silicosis were excluded from this table due to having no positive example in the test set.

### Binomial proportions estimated from extracted variables

The estimated proportions and their 95% confidence intervals created from the “Yes/No/Not recorded” and “Positive/Negative” extracted variables are given in Table 4.

**Table 4:** Binomial proportion estimate and 95% confidence interval (CI) using standard binary regression and MC-SIMEX model for binary variables created from extracted variables. Total patients with at least 1 dictation: N = 3237.

| Description | Count (N=3237) | Logistic regression estimate (95% CI) | MC-SIMEX model estimate (95% CI) |
| --- | --- | --- | --- |
| **Demographics** |  |  |  |
| Healthcare workers | 438 | 13.5%<br>(12.4, 14.8) | 2.48%<br>(2.02, 3.04) |
| **Tuberculosis Diagnosis** |  |  |  |
| Known TB exposure | 706 | 21.8%<br>(20.4, 23.3) | 16.8%<br>(15.3, 18.3) |
| Received BCG vaccination | 1316 | 40.7%<br>(39.0, 42.4) | 24.8%<br>(23.0, 26.7) |
| Performed a TST | 2279 | 70.4%<br>(68.8, 72.0) | 69.7%<br>(68.1, 71.3) |
| Received a positive TST interpretation | 2031 | 62.7%<br>(61.1, 64.4) | 61.9%<br>(60.2, 63.6) |
| Performed an IGRA | 296 | 9.14%<br>(8.20, 10.2) | 9.14%<br>(8.20, 10.2) |
| Received a positive IGRA interpretation | 301 | 9.30%<br>(8.35, 10.3) | 9.30%<br>(8.35, 10.3) |
| Diagnosed with active TB | 640 | 19.8%<br>(18.4, 21.2) | 18.2%<br>(16.8, 19.7) |
| Diagnosed with LTBI | 1473 | 45.5%<br>(43.8, 47.2) | 39.7%<br>(37.8, 41.6) |

| **Tuberculosis Medications** |  |  |  |
| --- | --- | --- | --- |
| Ever started on isoniazid | 1314 | 40.6%<br>(38.9, 42.3) | 45.6%<br>(43.6, 47.5) |
| Ever started on rifampin | 548 | 16.9%<br>(15.7, 18.3) | 17.6%<br>(16.3, 19.1) |
| Ever started on pyrazinamide | 349 | 10.8%<br>(9.76, 11.9) | 9.99%<br>(8.96, 11.1) |
| Ever started on ethambutol | 348 | 10.8%<br>(9.73, 11.9) | 9.36%<br>(8.32, 10.5) |
| Ever started on vitamin B6 | 986 | 30.5%<br>(28.9, 32.1) | 30.6%<br>(29.0, 32.2) |
| **Medical Conditions and Comorbidities\*** |  |  |  |
| Autoimmune conditions | 167 | 5.16%<br>(4.45, 5.98) | 0.259%<br>(0.175, 0.383) |
| Diabetes | 179 | 5.53%<br>(4.79, 6.37) | 0.358%<br>(0.247, 0.517) |
| Hematological malignancy | 71 | 2.19%<br>(1.74, 2.76) | 0.00625%<br>(0.00320, 0.0122) |
| Non-hematological malignancy | 140 | 4.32%<br>(3.68, 5.08) | 0.860%<br>(0.599, 1.23) |
| Renal failure | 65 | 2.01%<br>(1.58, 2.55) | 0.00895%<br>(0.00450, 0.0180) |
| Diagnosed with HIV | 175 | 5.41%<br>(4.68, 6.24) | 5.43%<br>(4.69, 6.26) |
| No relevant medical conditions/comorbidities** | 2569 | 79.4%<br>(77.9, 80.7) | 89.3%<br>(87.9, 90.6) |

MC-SIMEX: Misclassification Simulation Extraction; CI: Confidence interval; TB: Tuberculosis; BCG: Bacillus Calmette–Guérin; TST: Tuberculin sensitivity test; IGRA: Interferon gamma release assay; LTBI: Latent tuberculosis infection; HIV: Human immunodeficiency viruses
\*Patients that had undergone a transplant and patients diagnosed with silicosis were excluded from this table due to having no positive example in the test set.
\*\*Includes any patient with an extracted value of “No/Not recorded/Negative” for all medical conditions/comorbidities listed in the table.

After accounting for misclassification errors, the proportion of patients with an active TB diagnosis was 18.2% and the proportion of patients with an LTBI diagnosis was 39.7%. 69.7% of patients had performed a tuberculin sensitivity test and 61.9% of all patients had a positive result for the test. The proportions of patients who were ever started on isoniazid, rifampin or B6 were 45.6%, 17.6% and 30.6% percent, respectively.

## Discussion

To facilitate research on TB clinical epidemiology, diagnostics, clinical care and program implementation, quality improvement, and linkage for future therapeutics trials and biomarker studies, we developed a retrospective database of TB clinic patients using structured and unstructured EHR data. The cohort and database are unique in the transformation of unstructured data into structured variables using natural language rulesets with excellent performance when validated against manual chart abstraction. The rulesets are open access, and the database is accessible for research and open for collaboration with approval from local research ethics board.

The strength of the SMH-TB database comes from the inclusion of granular data, achieved by extracting it from unstructured sources using natural language processing. While the database contains standard structured data accessible in a wide variety of EHRs, a large and unique component of our data comes directly from unstructured dictated clinic notes, which contain a vast number of variables that can be used for a broad range of research topics, including, for example, clinical epidemiology and modelling studies. The NLP rulesets allow us to create granular patient-level variables from unstructured data accurately and efficiently, reducing the amount of time spent on manual abstraction to a minimum. Moreover, the large amount of unstructured raw data is a tremendous resource for evaluating and deploying machine learning and deep learning models capable of automatically extracting meaningful variables from clinical notes (27–29). Machine learning models and workflows can be developed to leverage the structured and extracted variables for predictive modeling and early warning systems (30–32). The breadth of data provided makes this a unique and powerful tool in both clinical and computational research.

The main limitations of the SMH-TB database include issues that arise from missing or incorrect data and the limited availability of data for certain variables leading to non-robust natural language rulesets. Data errors can be due to both human and algorithmic mistakes. Much of the burden of including relevant data in clinical dictations lies with the clinician attending the patient and dictating the note. In the absence of a standardized format, as was the case in the SMH TB clinic, variables may not be dictated in a manner that enables their capture by the NLP tools, or are not dictated at all. The creation of a shared set of guidelines and standard formatting for TB clinic dictations, containing all variables relevant to the database, will ensure all data required are captured with future database updates.

When the unstructured data undergoes information extraction, mislabeling of variables can occur due to certain rulesets having subpar performance. This issue is especially apparent for variables with scarce availability of labels. For example, in our validation dataset there were no patients with silicosis. The ruleset for classifying silicosis was adapted from other immunosuppressive conditions and expert knowledge in disease. While it is possible that such rulesets are overly confident in assigning a “No” label to patients even if they present with the condition in question, given the rarity of the event in the patient population it was not possible to provide further cases for perfection of refinement of the NLP ruleset. As such, we have indicated the metrics of our variables (Table 3), so that researchers can understand the limitations of the data with which they are working. The 200 charts sampled for ruleset refinement were consecutive patients from a set of clinic visits and may not have been sufficient for less common variables such as comorbidities. That is, further ruleset refinement will be needed with additional charts with purposive sampling of true positives of infrequent variables.

## Conclusion

In summary, here we describe the SMH-TB cohort and database which aim to be a resource for scientists who are conducting research into many facets of TB. The database is unique in that it contains highly granular socio-demographic and clinical patient data derived from structured and unstructured EHR data extracted using NLP rule sets. The validated rule sets are provided open access for use and the data base is intended to be available for collaborative studies.

## Data Availability

The validated NLP rulesets are publicly available for use from: https://github.com/mishra-lab/tb-nlp-rulesets. Data collected in SMH-TB contains sensitive patient information and as such, researchers interested in conducting TB-related research using the data are welcome to contact the corresponding author and submit a request. The study team welcomes collaboration and use of the database, and all external requests will be screened to ensure adequate data exists to enable a collaboration. The project will then undergo the approval process of the Research and Ethics Board (REB) of Unity Health Toronto. Data provided to researchers can either be the de-identified version of the SMH-TB database, or the full identifiable version, based on their research needs and REB approval.

## Data Availability

The validated NLP rulesets are publicly available for use from: https://github.com/mishra-lab/tbnlp-rulesets. Data collected in SMH-TB contains sensitive patient information and as such, researchers interested in conducting TB-related research using the data are welcome to contact the corresponding author and submit a request. The study team welcomes collaboration and use of the database, and all external requests will be screened to ensure adequate data exists to enable a collaboration. The project will then undergo the approval process of the Research and Ethics Board (REB) of Unity Health Toronto. Data provided to researchers can either be the de-identified version of the SMH-TB database, or the full identifiable version, based on their research needs and REB approval.

## Acknowledgements

We thank Dr. Natasha Sabur for supporting arbitration for rulesets; Julie Seemangal (TB Outpatient Clinic Co-Lead) and Grace Bezaliel for supporting verification of algorithms to classify patients seen in the TB clinic.

## Author Contributions

David Landsman – Data Curation, Formal Analysis, Investigation, Methodology, Software, Validation, Visualization, Writing – Original Draft Preparation, Writing – Review & Editing

Ahmed Abdelbasit – Data Curation, Investigation, Validation, Writing – Original Draft Preparation, Writing – Review & Editing

Christine Wang – Data Curation, Investigation, Validation, Writing – Review & Editing

Michael Guerzhoy – Investigation, Methodology, Software, Writing – Review & Editing

Ujash Joshi – Investigation, Methodology, Software, Writing – Review & Editing

Shaun Mathew – Investigation, Methodology, Software, Writing – Review & Editing

Chloe Pou-Prom – Investigation, Methodology, Software, Writing – Review & Editing

David Dai – Investigation, Methodology, Software, Writing – Review & Editing

Victoria Pequegnat – Data Curation, Resources, Validation, Writing – Review & Editing

Joshua Murray – Investigation, Methodology, Software, Writing – Review & Editing

Kamalprit Chokar – Data Curation, Writing – Review & Editing

Michaelia Banning – Project Administration, Writing – Review & Editing

Muhammad Mamdani – Project Administration, Writing – Review & Editing

Sharmistha Mishra – Conceptualization, Data Curation, Funding Acquisition, Investigation, Methodology, Project Administration, Resources, Supervision, Validation, Writing – Review & Editing

Jane Batt – Conceptualization, Data Curation, Funding Acquisition, Investigation, Methodology, Project Administration, Resources, Supervision, Validation, Writing – Review & Editing

